# Noninvasive electrocardiographic imaging assessment of intraventricular synchrony for conduction system pacing device: a novel algorythm to assess intraventricular synchrony

**DOI:** 10.1101/2024.08.14.24311086

**Authors:** Ivan Eltsov, Alvise Del Monte, Luigi Pannone, Ingrid Overeinder, Domenico Della Rocca, Roberto Scacciavillani, Frederik H Verbrugge, Qingguo Zeng, Gezim Bala, Andrea Maria Paparella, Giacomo Talevi, Erwin Stroker, Juan Sieira, Ali Gharaviri, Andrea Sarkozy, Gian-Battista Chierchia, Mark La Meir, Carlo de Asmundis, Alexandre Almorad

## Abstract

**Background:** Left Bundle branch area pacing has become the procedure of choice for various indications including atrioventricular block and considered to be a physiologic modality of pacing compared to RV apex pacing.

**Objectives:** The purpose of this study was to assess ventricular activation and synchrony in patients with LBBAP device using ECG imaging (ECGI).

**Methods:** 25 consecutive patients underwent an LBBAP device implantation have been included in the study. ECG and ECGI analysis have been performed the day after implantation. Native and paced QRS, LVAT, RVAT and V1AD were calculated using ECG. TVACT, LVACT, LVACTi, RVACT, RVACTi and IVDS were calculated based on ECGI. All patients have been followed up for 12 months.

**Results:** All patients were divided in 2 groups (wide and narrow QRS) based on intrinsic ECG and then based on paced ECG QRS.

For initially narrow QRS group, activation time and synchrony during pacing was comparable to native. In wide QRS group these parameters were significantly improved.

At paced rhythm analysis, classic ECG LBBAP parameters (paced QRS and LVAT) were not sufficient to properly evaluate the ventricular activation for paced rhythm. Discordance between ECG and ECGI analysys was identified in 25 patients. Two additional 12 lead ECG parameters predicting the ECGI measurements were found - V1AD and dRVAT. Follow up showed stable values of ejection fraction, paced QRS and pacing parameters.

**Conclusions:** ECG imaging can bring a significant value into assessing the efficacy of new pacing modalities and provide more data for precise determination of implantation outcomes, including detailed activation assessment and comparison to intrinsic conduction. Key ECGI values confirming proper ventricular activation have been defined and correlated with 12 lead ECG parameters to predict ventricle activation from ECG only.

## 1. Introduction

In the last few years, the left bundle branch area pacing (LBBAP) technique has become the procedure of choice for patients with a structurally normal heart requiring permanent ventricular pacing (e.g., atrioventricular block and persistent atrial fibrillation) (1). Conduction-system pacing (CSP), especially high-septal pacing (HSP), avoids pacemaker syndrome and preserves the ejection fraction during permanent ventricular pacing. The most common physiological pacing technique is His bundle pacing (HBP); however, the main advantage of LBBAP is that similar physiological ventricular pacing can be achieved with higher success rates and stable pacing values at follow-up.

Despite the huge interest in the LBBAP technique, published data remain scarce and definitions and distinctions between these modalities are not uniform, varying between different researchers (2). Patients with a wide QRS complex could benefit from the implantation with improvement in the ejection fraction (3). LBBAP shortens the QRS complex significantly compared with right ventricle (RV) pacing (4) and full correction of a left bundle branch block (LBBB) can more often and more easily be achieved compared with HBP (2).

In the recent MELOS study, Jastrzebsky et al. concluded that LBBAP is a feasible technique to treat bradyarrhythmias; however, the success rate must be improved and clinical outcomes investigated in randomized trials (5).

The main keys to success in LBBAP implantation are the implantation site, intraventricular synchrony and pacing parameters. Currently, the standard way to assess activation during and after LBBAP implantation is surface electrocardiogram (ECG) and conduction system potentials. This requires not only solid ECG interpretation skills of the operator but often a second person at the recording system to analyze the activation. The same limitation may occur in the postprocedural follow-up.

Cheng et al. have recently shown that intraventricular synchrony can be achieved not only in pure left bundle branch pacing (LBBP) groups but also in other LBBAP modalities (6).

We hypothesized that noninvasive electrocardiographic imaging (ECGI) could be beneficial for intra- and post-implantation evaluation and optimization of LBBAP device therapy by precise visualization of ventricular activation and synchrony. This noninvasive high-fidelity technique allows the generation of activation maps and timings (7). Other researchers have successfully used this technique for ventricular activation assessment after cardiac resynchronization therapy (CRT) device implantation, concluding that it provides reliable ventricular activation data and may be a useful adjunct to guide left ventricle lead implants and to perform postimplant CRT optimization. (7, 8). Another advantage of this technique is that it overcomes ECG interpretation issues, which may occur while dealing with patients with abnormal anatomy and/or suboptimal ECG patch placement.

## 2. Materials and Methods

### Study design

This single-center prospective study included 25 patients who underwent LBBAP device implantation followed by ECGI evaluation. The study complies with the Declaration of Helsinki and was approved by the local ethics committees; informed consent was obtained from the subjects before inclusion in the study. All data were collected and updated in the registry of the Universitair Ziekenhuis Brussel and approved by the institutional ethics committee.

### Study endpoints

The primary endpoint was the achievement of the proper conduction system pacing using LBBAP technique following current guidelines (1). Other endpoints were ECGI assessment of intraventricular synchrony in different patient cohorts (narrow and wide QRS) and evaluation of the differences between intrinsic and paced rhythms.

### Patient population

Twenty-five consecutive patients who underwent LBBAP implantation were analyzed and included in the study. The inclusion criteria were: 1) a confirmed indication for ventricular pacing and 2) stable leads position and parameters the day after implantation.

Exclusion criteria were the presence of structural heart disease and severe heart failure requiring CRT.

### LBBAP implantation procedure

Two different LBBAP implantation techniques were used. A stylet-driven lead (Solia S60, Biotronik, Germany) and a non-stylet-driven lead (SelectSecure 3830, Medtronic, USA) implantation with compatible sheath. Both techniques are well described in the literature; (9, 10, 11) the main procedural difference was that the use of leads with stylet allows the assessment of conduction system potentials and the morphology of paced QRS while screwing the lead into the ventricular septum. (9).

The main steps of the procedure can be summarized as follows: (1) transvenous access; (2) intraseptal placement of the pacing lead into the left ventricle (LV) septal subendocardium in the left bundle branch (LBB) area; (3) confirmation of LBB capture; (4) placement of atrial connection leads in the pacemaker (PM), and (5) pocket closure. Procedures were performed either under general or local anesthesia. The day following implantation, the lead position was controlled via chest imaging, and complete device interrogation was performed to confirm stable pacemaker parameters.

Each implantation was assessed using standard 12-lead ECGs to measure classic LBBAP values: QRS, left and right ventricle activation times (LVAT and RVAT), and V1-V6 activation delay (V1AD). All values were measured for intrinsic and paced rhythms.

### Electrocardiographic imaging procedure

The ECGI procedure was performed one day after implantation, before discharge. A mapping Vest (CardioInsight, Medtronic Inc, MN, USA) was applied to the patient’s chest and signal acquisition was started. Then, the PM configuration was modified to record both intrinsic and paced rhythms and the corresponding bookmarks were created in the mapping system. Patients then underwent a chest CT scan, which was segmented to align 252 surface electrodes with the epicardial shell (Figure 1).

**Figure 1.**
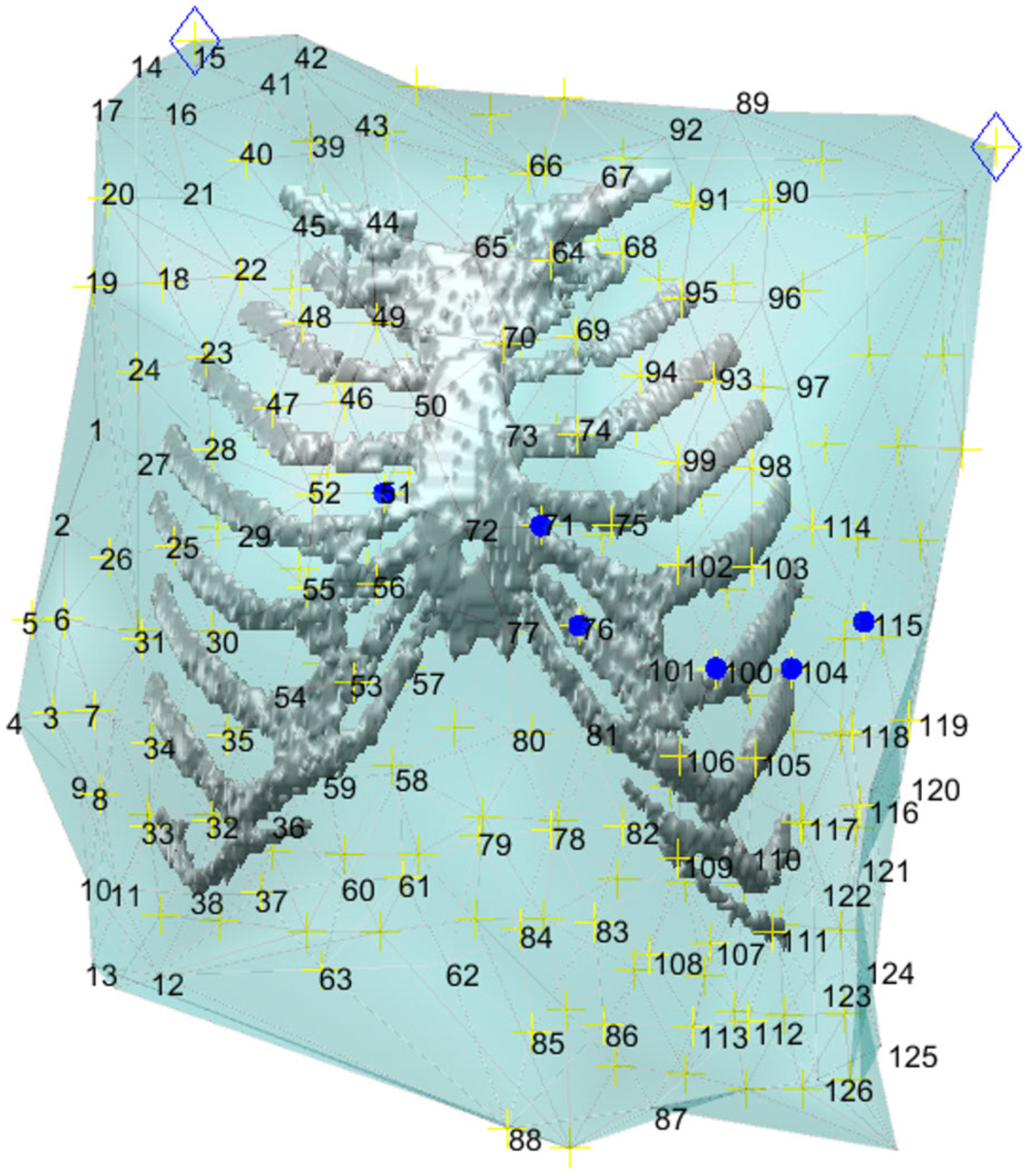
ECG and ECGI acquisition differences. Chest CT scan with thoracic cage and vest electrode markers (yellow crosses). Blue dots are the reference positions of classic V leads Later, activation and propagation maps were created. In our study, we performed single-beat mapping of the QRS complex for baseline (intrinsic) and paced rhythms (Figures 2 A and B).

### ECGI analysis

Different ventricle activation parameters of intrinsic conduction were measured, namely: 1) Total ventricle activation time (TVACT) - the time interval between the first negative dv/dt of the earliest ventricular unipolar electrogram and the last dv/dt of the latest ventricular unipolar electrogram (Figure 2); 2) left ventricle activation time (LVACT) - the time interval between the pacing spike and the latest ventricular unipolar electrogram of the left ventricle (Figure 3); 3) right ventricle activation time (RVACT) – the time interval between the pacing spike and the latest ventricular unipolar electrogram on the right ventricle (Figure 3). A dynamic potential map was used to identify all the earliest and latest activation points on the map, and directional activation maps were used to measure time intervals; 4) Intraventricular dyssynchrony (IVDS) was calculated as the difference between RVACT and LVACT.

**Figure 2.**
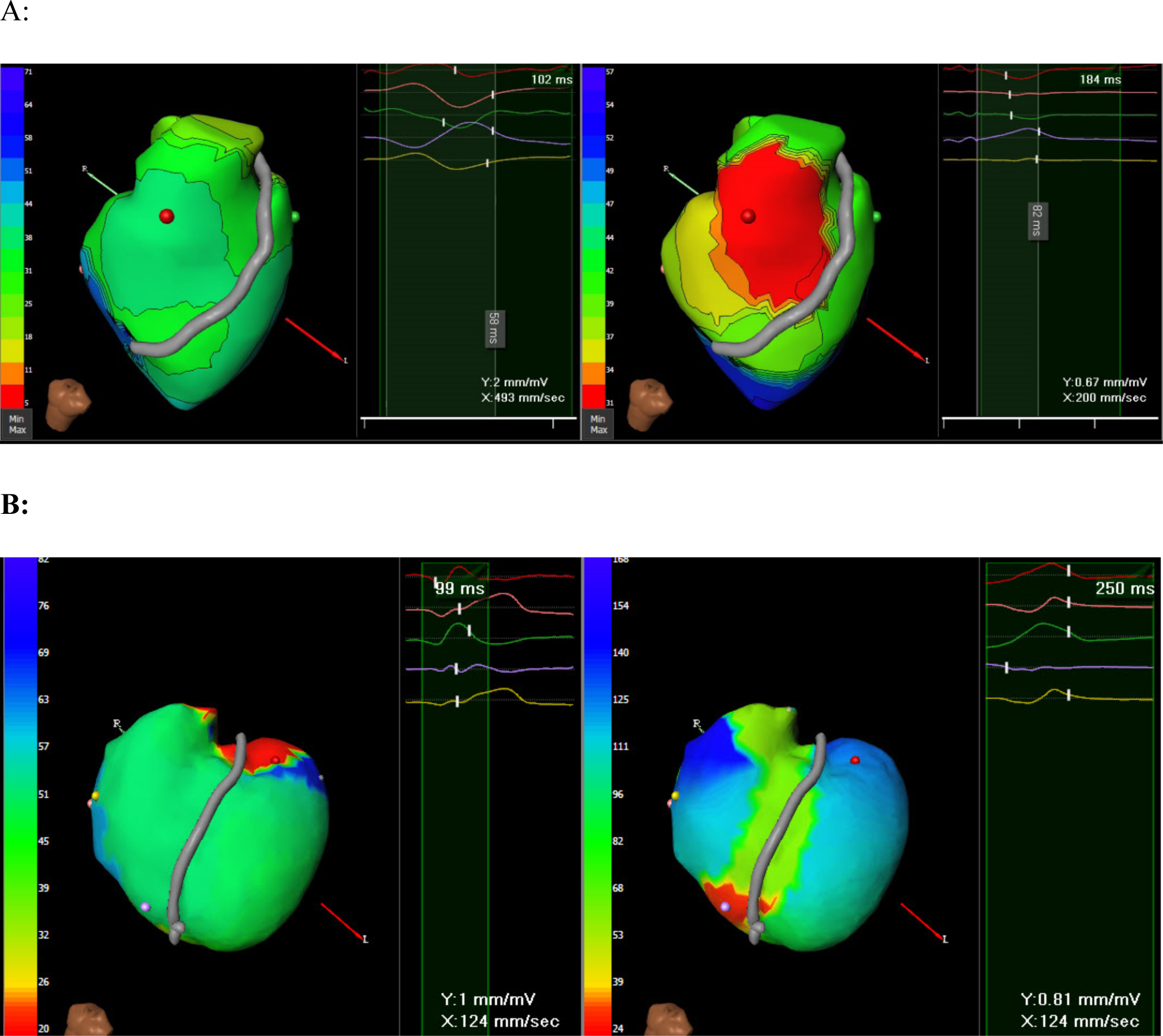
Intrinsic and paced activation maps of the same patient A: narrow intrinsic QRS, B: wide intrinsic QRS. Red areas represent the earliest activation site and blue the latest.

**Figure 3.**
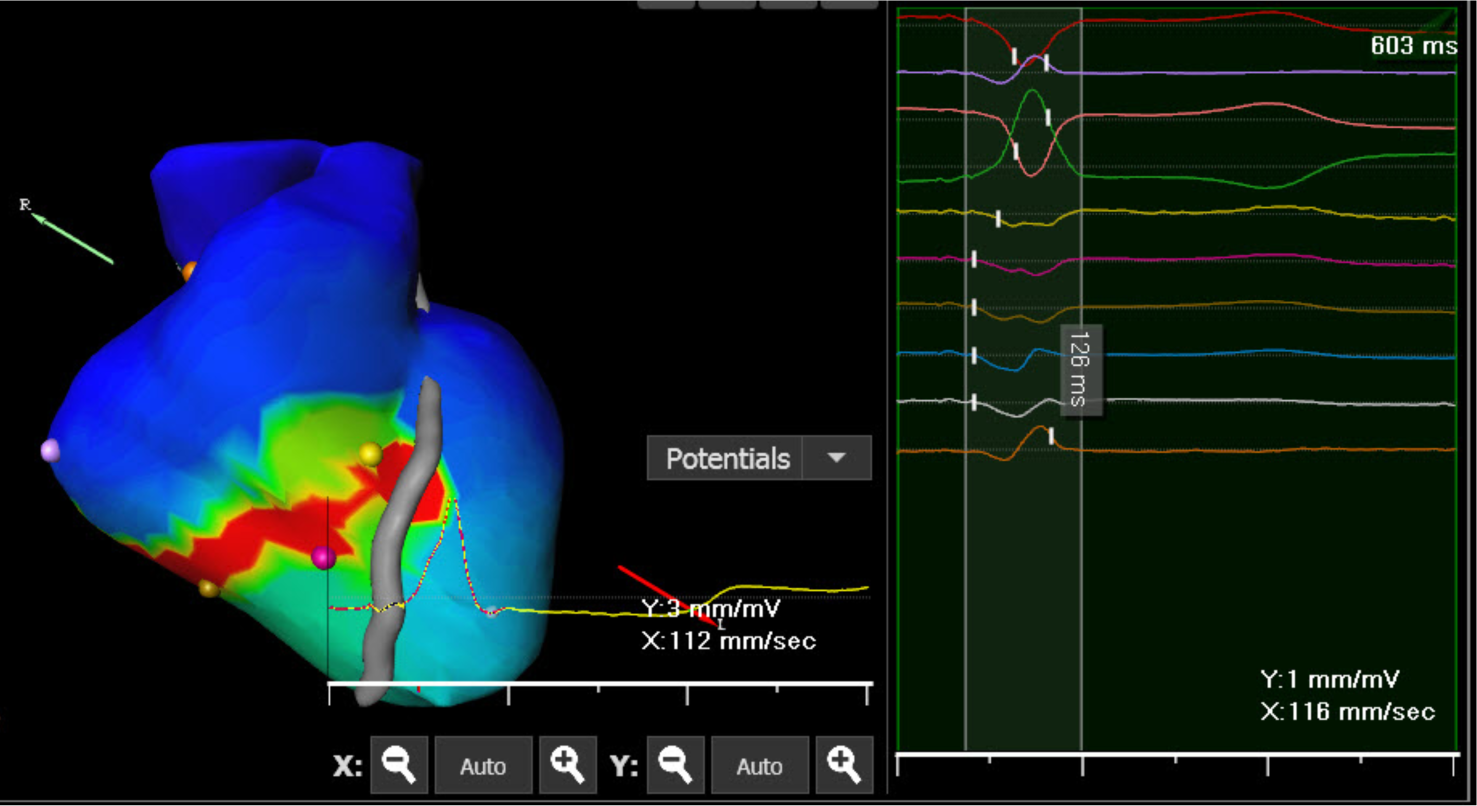
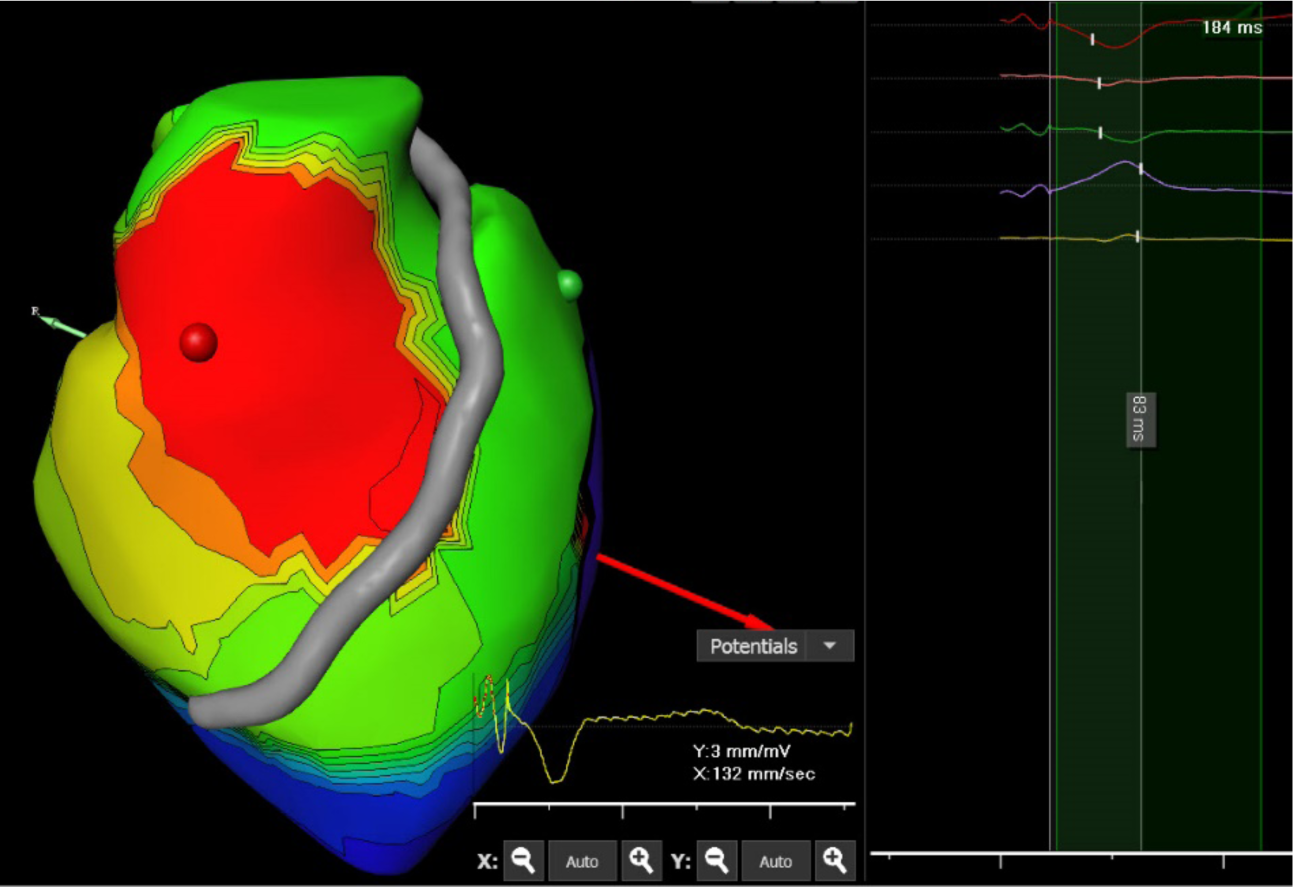
TVACT, LVACT, and RVACT measurement on the ECGI Activation map. A: TVACT measurements for paced rhythm – white caliper (126 ms) measuring from the first deflection of the earliest activation spot to the last deflection of the latest activation spot. **B:** LVACT and RVACT measurements for almost concomitant activation (left ventricle activated slightly earlier). The white caliper measures RVACT (83 ms) from the pacing spike to the steepest negative dv/dt at the latest RV activation point.

For paced rhythm, the pacing spike was used as the starting measurement point for all intervals to ensure the consistency for comparison between ECG and ECGI timings.

Subsequently, both ECG and ECGI results were compared to identify potential discordance within different groups. TVACT values > 130 ms (long TVACT) were defined as a confirmation criterion of wide-paced QRS after LBBAP implantation, while TVACT values ≤ 130 ms (short TVACT) were used as a confirmation of narrow-paced QRS.

Every ECG-based value has an ECGI-based equivalent; LVAT – LVACT, dRVAT – RVACT, IVDS – V1AD. In addition, the difference between LV activation measured using two different techniques was analyzed. LBBAP pacing was considered “optimal” if the TVACT value was < 130 ms and IVDS positive or around zero.

All ECG measurements were also repeated for intrinsic and paced rhythm in all groups to identify clinical pacing outcomes.

### Statistical analysis

The analysis was performed using R software version 3.6.2 (R Foundation for Statistical Computing, Vienna, Austria).

All variables were tested for normality with the Shapiro–Wilk test. Normally distributed variables were described as mean ± standard deviation and groups were compared through ANOVA, paired or unpaired t-test as appropriate. Non-normally distributed variables were described as median (interquartile range) and compared using the Mann–Whitney or Wilcoxon signed-rank test, as appropriate. The categorical variables were described as frequencies (percentages) and compared by the χ2 or Fisher’s exact test, as appropriate. A *P*-value < 0.05 was considered statistically significant.

## 3. Results

### Study population characteristics

Twenty-five consecutive patients who underwent LBBAP implantation met the inclusion criteria and were subjected to an ECGI procedure the day following implantation. All patients showed correct PM lead position via chest x-ray and stable sensing and pacing parameters during device interrogation. Twelve (48%) procedures were performed using stylet-driven leads from Biotronik and Medtronic lumenless leads were used in the remaining 13 patients (52%). Based on a preprocedural 12-lead ECG, patients were divided into two groups: narrow QRS and wide QRS. Patient characteristics are summarized in Table 1.

**Table 1.** Patient characteristics.

|  | **Narrow QRS**<br>**(N=16)** | **Wide QRS**<br>**(N=9)** | **Overall**<br>**(N=25)** | ***P*-value** |
| --- | --- | --- | --- | --- |
| **Age (years)** | 66.1 ± 16.5 | 73.7 ± 16.0 | 68.8 ± 16.5 | 0.28 |
| **Sex (male)** | 7 (43.8%) | 9 (100%) | 16 (64.0%) | 0.008 |
| **BMI (Kg/m2)** | 30.0 ± 5.0 | 29.1 ± 5.0 | 29.7 ± 4.9 | 0.64 |
| **Baseline ECG morphology** |  |  |  | 0.003 |
| **LBB morphology** | 7 (43.8%) | 2 (22.8%) | 9 (36.0%) |  |
| **RBB morphology** | 2 (12.5%) | 7 (77.8%) | 9 (36.0%) |  |
| **Normal** | 7 (43.8%) | 0 (0.0%) | 7 (28.0%) |  |
| **Pre-implantation QRS duration (ms)** | 95.1 ± 12.2 | 153.4 ± 16.9 | 112.9 ± 30.5 | <0.001 |
| **Ejection Fraction (%)** | 55.3 ± 5.0 | 53.9 ± 4.2 | 54.8 ± 4.7 | 0.48 |
| **PR (ms)** | 214.2 ± 76.7 | 191.2 ± 56.1 | 207.8 ± 70.8 | 0.55 |
| **QTc (ms)** | 434.2 ± 32.0 | 460.3 ± 41.8 | 443.6 ± 37.2 | 0.09 |
| **SDL vs. LLL** | 8/8 | 5/4 | 12/13 | 1.0 |
| **Sick Sinus Syndrome** | 13 (81.2%) | 5 (55.6%) | 18 (72.0%) | 0.21 |
| **AV block** | 5 (31.2%) | 5 (55.6%) | 10 (40.0%) | 0.39 |
| **Intraventricular Conduction Delay** | 0 (0.0%) | 1 (11.1%) | 1 (4.0%) | 0.36 |

BMI – Body mass index, LBB – Left Bundle Branch Block, RBB – Right Bundle Branch Block, SDL – Stylet-driven lead, LLL – Lumenless lead, AV – atrioventricular.

At the end of the procedure, based on the ECG assessment, all patients were divided into two groups: pQRS < 130 ms and ≥ 130 ms. ECG measurements for the two pQRS groups are summarized in Table 2.

**Table 2.** ECG measurements for LBBAP pacing.

|  | **pQRS < 130 ms**<br>**(N=11)** | **pQRS ≥ 130 ms**<br>**(N=14)** | **Total (N=25)** | ***P*-value** |
| --- | --- | --- | --- | --- |
| **pQRS** | 117.5 ± 6.2 | 138.6 ± 9.5 | 129.3 ± 13.4 | <0.001 |
| **LVAT** | 60.5 ± 8.6 | 77.1 ± 20.5 | 69.8 ± 18.2 | 0.019 |
| **RVAT** | 73.8 ± 20.2 | 98.0 ± 26.8 | 87.4 ± 26.6 | 0.021 |
| **V1AD** | 16.1 ± 17.3 | 20.2 ± 37.7 | 18.4 ± 30.0 | 0.74 |

pQRS – paced QRS, LVAT – Left Ventricle Activation Time (ECG), RVAT – Right Ventricle activation time (ECG), V1AD – V1 Activation delay (ECG)

The comparison of ECGI data between pQRS < 130 ms and ≥ 130 ms is summarized in Table 3.

**Table 3.**
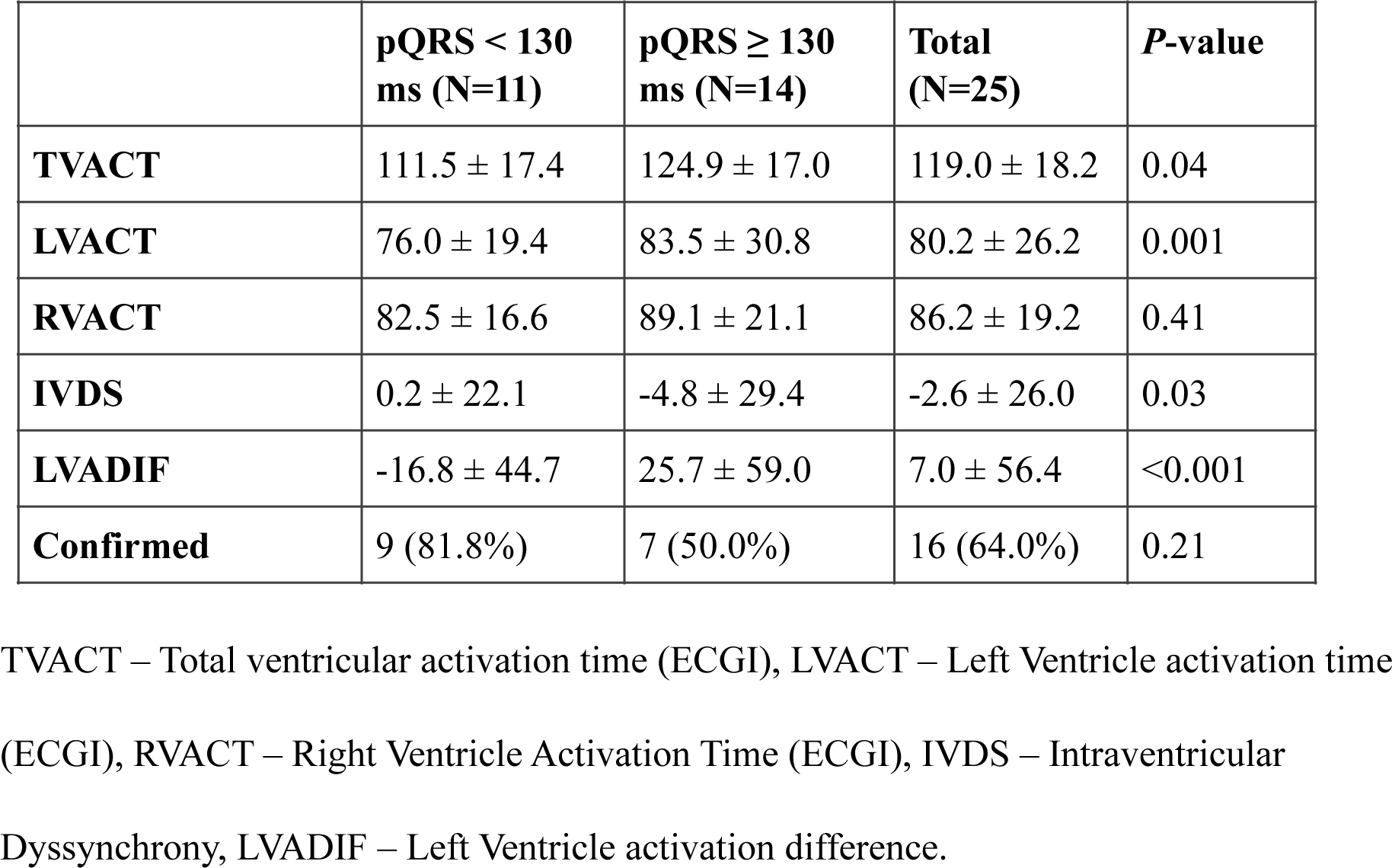
Noninvasive ECGI analysis for pQRS groups.

|  | **pQRS < 130**<br>**ms (N=11)** | **pQRS ≥ 130**<br>**ms (N=14)** | **Total**<br>**(N=25)** | ***P*-value** |
| --- | --- | --- | --- | --- |
| **TVACT** | 111.5 ± 17.4 | 124.9 ± 17.0 | 119.0 ± 18.2 | 0.04 |
| **LVACT** | 76.0 ± 19.4 | 83.5 ± 30.8 | 80.2 ± 26.2 | 0.001 |
| **RVACT** | 82.5 ± 16.6 | 89.1 ± 21.1 | 86.2 ± 19.2 | 0.41 |
| **IVDS** | 0.2 ± 22.1 | -4.8 ± 29.4 | -2.6 ± 26.0 | 0.03 |
| **LVADIF** | -16.8 ± 44.7 | 25.7 ± 59.0 | 7.0 ± 56.4 | <0.001 |
| **Confirmed** | 9 (81.8%) | 7 (50.0%) | 16 (64.0%) | 0.21 |

TVACT – Total ventricular activation time (ECGI), LVACT – Left Ventricle activation time (ECGI), RVACT – Right Ventricle Activation Time (ECGI), IVDS – Intraventricular Dyssynchrony, LVADIF – Left Ventricle activation difference.

Out of 14 patients with pQRS ≥ 130 ms, ECGI analysis confirmed only 7 LBBAP as suboptimal (TVACT ≥ 130 ms). Two patients from the narrow pQRS group (pQRS < 130 ms) had TVACT ≥ 130 ms.

The examples of concomitant (short TVACT) ventricular activation in the wide pQRS group and “Right first” activation (long TVACT), despite good classic ECG values, are shown in Figure 4.

**Figure 4.**
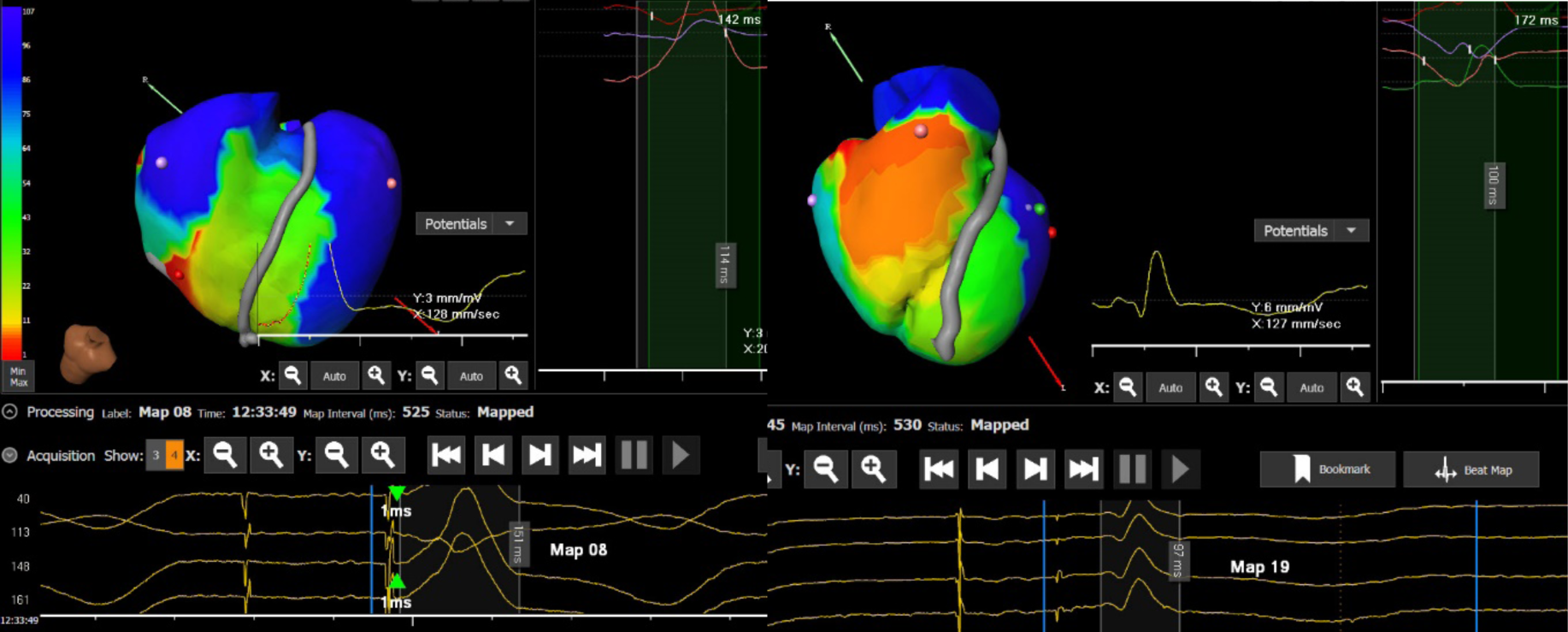
Left panel – concomitant activation of both ventricles in a wide pQRS patient. Right panel – “right first” activation of a patient with good classic LBBAP ECG measurements.

Analysis of the entire wide-paced QRS group showed that confirmed wide pQRS LBBAP are associated with higher TVACT and negative IVDS – both ECG and ECGI parameters are summarized in Table 4.

**Table 4.**
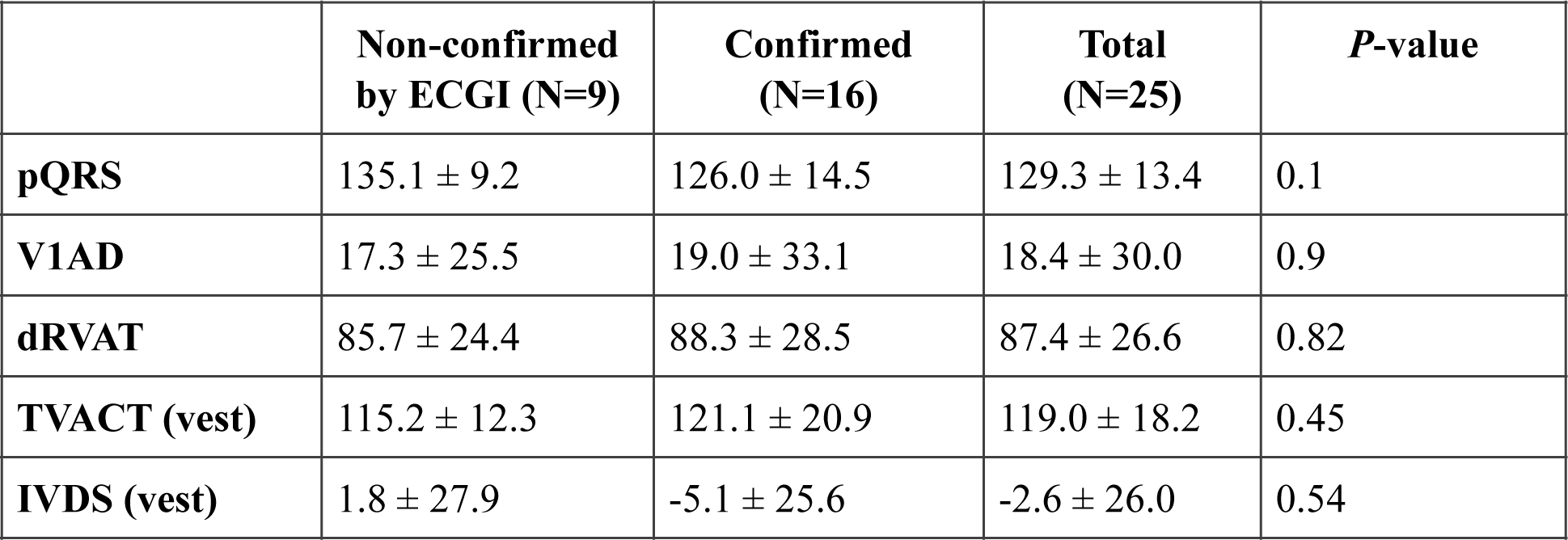
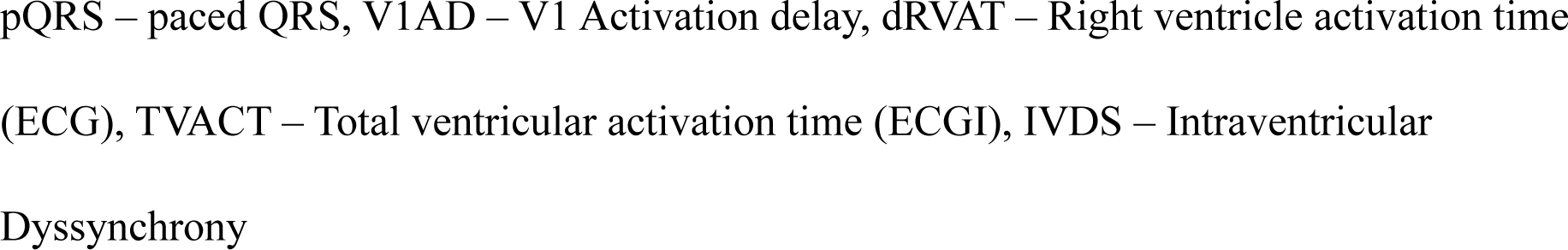
ECGI versus 12-lead ECG parameters in non-confirmed wide-paced QRS patients.

|  | **Non-confirmed**<br>**by ECGI (N=9)** | **Confirmed**<br>**(N=16)** | **Total**<br>**(N=25)** | ***P*-value** |
| --- | --- | --- | --- | --- |
| **pQRS** | 135.1 ± 9.2 | 126.0 ± 14.5 | 129.3 ± 13.4 | 0.1 |
| **V1AD** | 17.3 ± 25.5 | 19.0 ± 33.1 | 18.4 ± 30.0 | 0.9 |
| **dRVAT** | 85.7 ± 24.4 | 88.3 ± 28.5 | 87.4 ± 26.6 | 0.82 |
| **TVACT (vest)** | 115.2 ± 12.3 | 121.1 ± 20.9 | 119.0 ± 18.2 | 0.45 |
| **IVDS (vest)** | 1.8 ± 27.9 | -5.1 ± 25.6 | -2.6 ± 26.0 | 0.54 |

pQRS – paced QRS, V1AD – V1 Activation delay, dRVAT – Right ventricle activation time (ECG), TVACT – Total ventricular activation time (ECGI), IVDS – Intraventricular Dyssynchrony

Based on the results shown in Table 4, two ECG values (V1AD and dRVAT) could predict TVACT and IVDS values. Figure 5 shows the workflow defined from these data, which identifies ECGI-guided physiological pacing based on ECG analysis alone.

**Figure 5.**
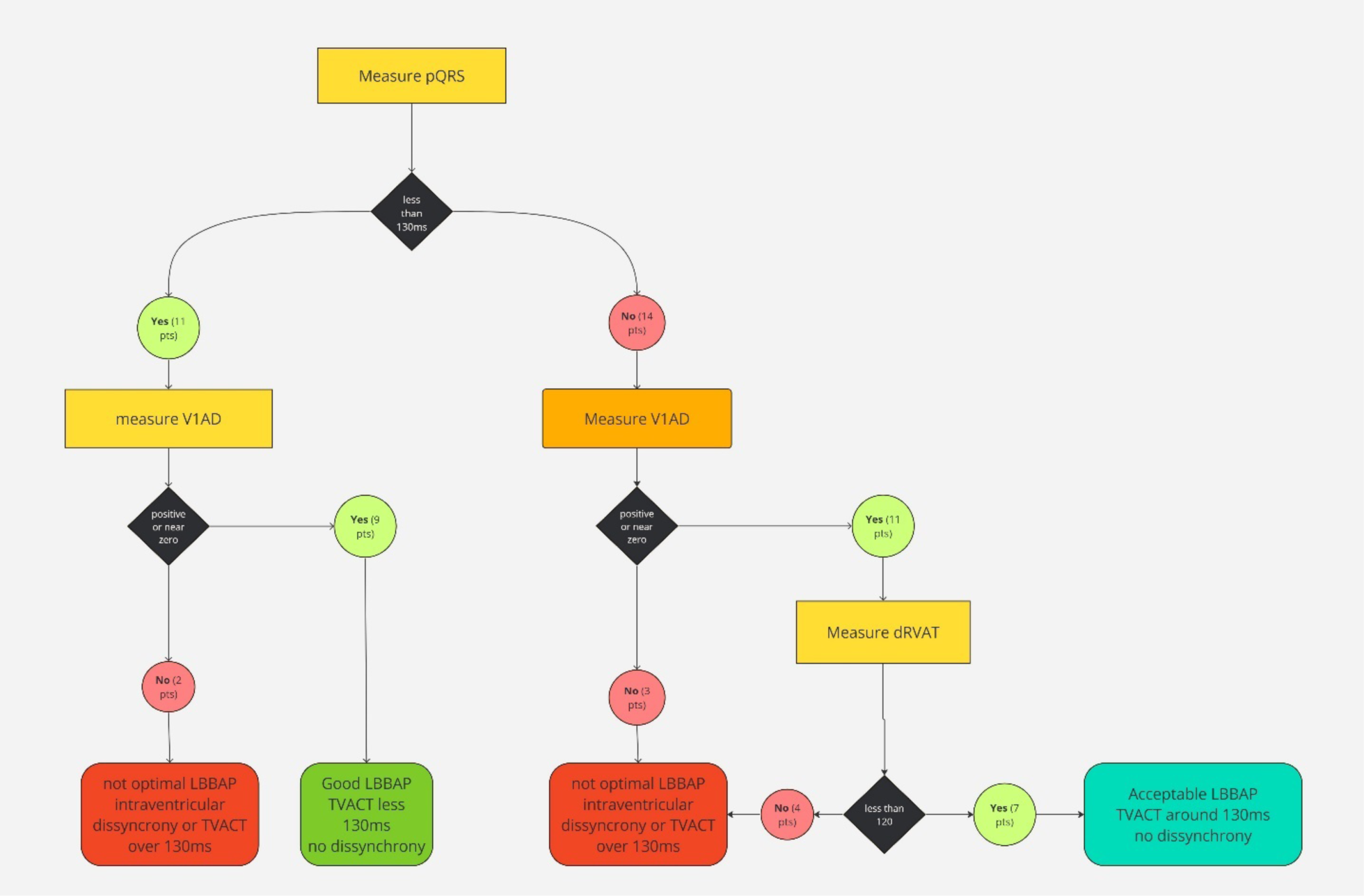
Suggested paced-rhythm ECG analysis workflow.

Finally, we performed an ECG and ECGI analysis of the intrinsic rhythm in patients with baseline narrow vs. wide QRS to compare the results achieved with LBBAP pacing (Table 5).

**Table 5.** Intrinsic vs. paced ECG and ECGI measurements.

|  | **Narrow QRS**<br>**(N=16)** | **Wide QRS**<br>**(N=9)** | **Overall**<br>**(N=25)** | ***P*-value** |
| --- | --- | --- | --- | --- |
| **Pre-implantation QRS duration (ms)** | 95.1 ± 12.2 | 153.4 ± 16.9 | 112.9 ± 30.5 | <0.001 |
| **pQRS (ms)** | 125.8 ± 13.0 | 135.6 ± 12.4 | 129.3 ± 13.4 | 0.08 |
| **TVACT (ms)** | 113.9 ± 17.5 | 128.0 ± 16.5 | 119.0 ± 18.2 | 0.06 |
| **RVACT (ms)** | 84.8 ± 22.6 | 88.8 ± 11.6 | 86.2 ± 19.2 | 0.62 |
| **RVACTi** | 30.6 ± 21.6 | 78.8 ± 41.8 | 47.9 ± 37.8 | <0.001 |
| **LVACT (ms)** | 74.5 ± 27.5 | 90.3 ± 21.4 | 80.2 ± 26.2 | 0.15 |
| **LVACTi** | 34.1 ± 13.0 | 60.3 ± 30.4 | 43.5 ± 24.1 | 0.006 |
| **IVDS** | -2.2 ± 24.4 | -3.2 ± 30.3 | -2.6 ± 26.0 | 0.93 |
| **IVDSi** | -3.5 ± 24.3 | 18.4 ± 68.0 | 4.4 ± 45.0 | 0.25 |
| **IVDS DIF** | -1.2 ± 40.2 | 21.7 ± 78.3 | 7.0 ± 56.4 | 0.34 |

pQRS – paced QRS, TVACT – Total ventricular activation time (ECGI), RVACT – Right ventricle activation time (ECGI), RVACTi – Intrinsic RVACT (ECGI), LVACT – Left ventricular activation Time (ECGI), LVACTi – intrinsic LVACT (ECGI), IVDS – intraventricular dyssynchrony, IVDSi – intrinsic IVDS, IVDS DIF – difference between IVDS and IVDSi.

In the narrow QRS group, the results show that intrinsic and paced IVDS were not significantly different, signifying that LBBAP activated the ventricles in almost exactly the same way as the native conduction system (Figure 6).

**Figure 6.**
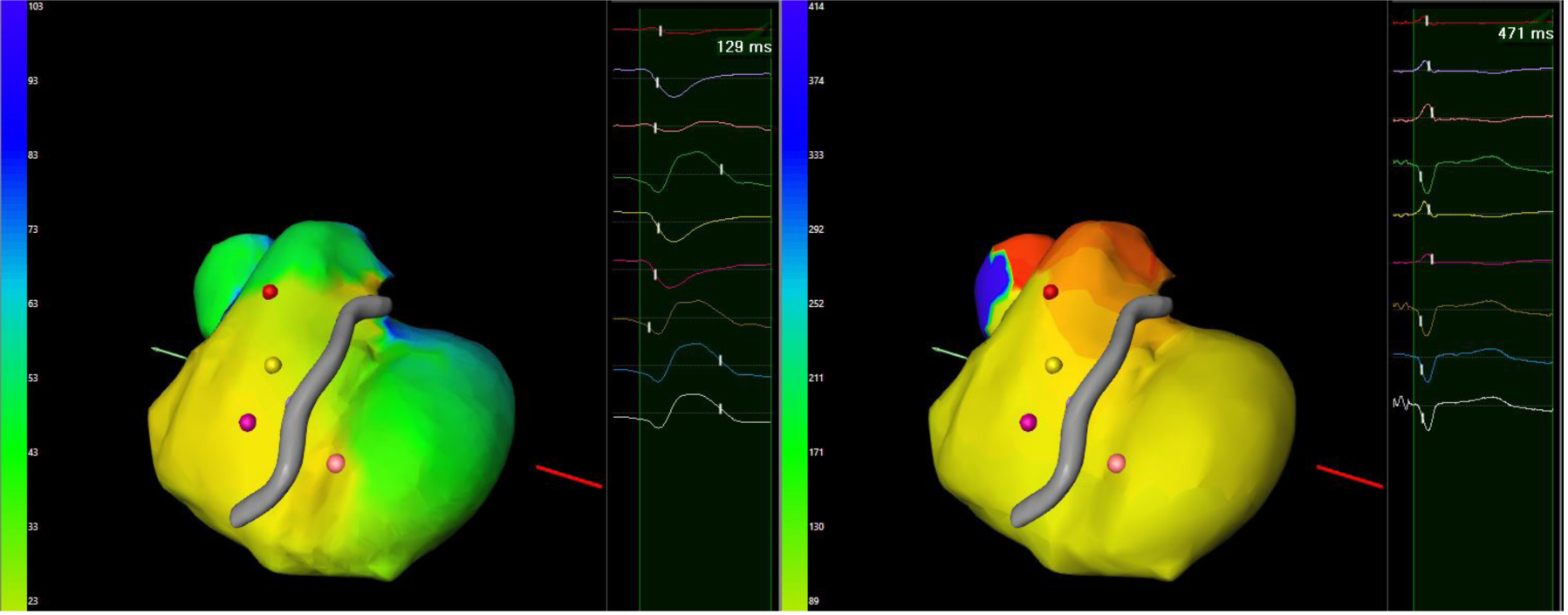
Activation maps of native and paced Rhythms in Patients with narrow intrinsic QRS.

However, in the wide QRS group, we found a significant improvement in intraventricular synchrony, and while the intrinsic rhythm was, as expected, not synchronous, the pQRS in this group was as good as in the narrow QRS group (Figure 7). This allowed us to conclude that, in the wide QRS group, the results of LBBAP were not different from the narrow QRS group based on pQRS and TVACT, RVACT, and IVDS.

**Figure 7.**
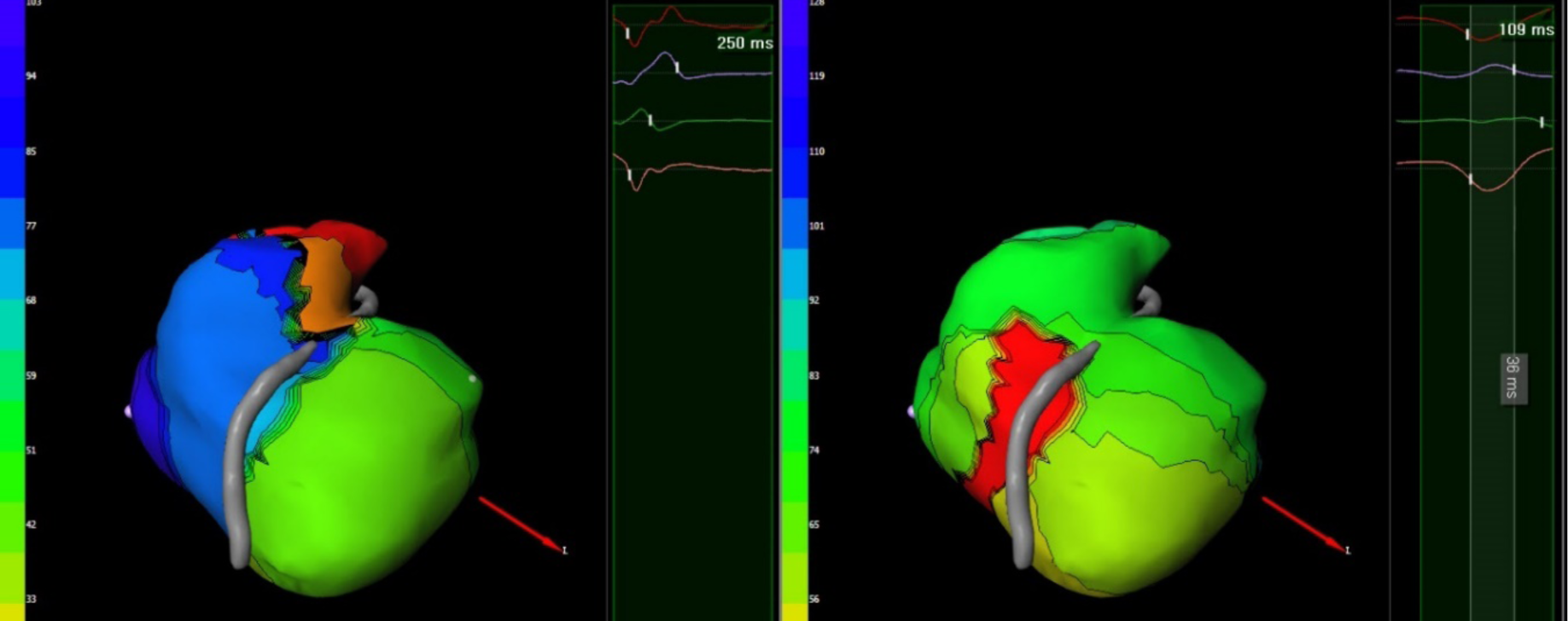
Improved ventricular activation in native wide QRS patients (Right bundle branch block-RBBB). Left: Intrinsic activation map. Right: Paced activation map. Red denotes the earliest activation point (pacing electrode).

12 month follow up didn’t show any worsening of EF, paced QRS or pacing parameters, follow up data is summarised in the Table 6.

**Table 6.** Twelve-months follow-up results.

|  | Baseline (N=25) | 12-months Follow-up (N=25) | Total (N=25) | p value |
| --- | --- | --- | --- | --- |
| **LVEF** | 54.8 (4.7) | 57.0 (4.6) | 55.9 (4.7) | 0.099 |
| **impedance** | 524.0 (103.5) | 493.3 (120.7) | 508.6 (112.3) | 0.339 |
| **threshold** | 0.6 (0.2) | 0.7 (0.4) | 0.7 (0.3) | 0.078 |
| **amplitude** | 12.7 (3.7) | 12.2 (5.2) | 12.5 (4.4) | 0.685 |

## 4. Discussion

### Feasibility of activation visualization via ECG imaging

Several research groups have already demonstrated the feasibility of using ECGI system to analyze different paced rhythms (RV, CRT, LBBAP) and compare them visually to the intrinsic rhythm, even using this data to optimize pacing modalities (CRT). (6,7,8,12-14), total activation time has been described by other investigators (15, 16).

In our study, in addition to visualization of activation during intrinsic and paced rhythms, activation times based on ECGI data were measured and compared with classic LBBAP assessment. We believe that the use of 252 electrodes around the torso during ECGI mapping allows us to obtain much more detail of the activation. Together with CT scan details, it allows a better analysis of activation than classic 12-lead ECG, which can lead to missing some information due to anatomical differences and variable patch positioning.

### True LBBAP confirmation

To achieve our primary endpoint, we analyzed the results of classic 12-lead ECG measurements and an ECGI study performed on the same day (one day after device implantation). As ECGI measurements were conducted slightly differently (from spike to latest deflection) and because the ECGI system can visualize more data, as expected, the activation time values did not match entirely.

Individual activation times (LVACT and RVACT) mostly followed the trends of LVAT and RVAT with a difference of approximately 10 ms; however, the intraventricular synchrony measured classically and using ECGI presented greater discordance. This might be related to the fact that classic 12-lead ECG cannot see the “left-most” activation and is limited to the V6 electrode position. ECGI is projecting 360 degrees of activation onto a precise 3D model, affording greater accuracy (7, 8).

In our study, we have used LVACT and RVACT values measured from spike until the last left or right deflection on ventricular activation and the difference between these values as a criterion for confirmation of “true LBBAP” implantation. The rationale for this is that LV and RV activation times measured with ECGI were higher than those measured via ECG because instead of measuring up to the R wave in V6 or V1, it measured until the last deflection.

We hypothesized that optimal LBBAP should be confirmed by two main ECGI parameters, TVACT and IVDS, as they most accurately reflect ventricle activation time and synchrony between LV and RV. After analysis, we found that physiological activation correlated with TVACT < 130 ms and IVDS positive (LV first) or around zero (concomitant activation).

With these parameters as control values, we reanalyzed our data and discovered that, in the short pQRS group, two patients did not have physiological ventricle activation despite good ECG parameters and, in the long pQRS group, seven patients had good ventricle activation.

Even though ECGI provides an added value to LBBAP implantation assessment, the procedure is associated with increased radiation exposure (CT scan) and extra cost (17).

Therefore, we performed an analysis to determine which 12-lead ECG measurements could predict similar results.

Our analysis demonstrated that there were two ECG values (V1AD and dRVAT) that best correlate with our results and can theoretically predict TVACT and IVDS values.

As expected, V1AD is the best predictor of IVDS as it measures the same process using different techniques. Thus, a negative V1AD is a discrimination factor, indicating that the right ventricle activates first. In our study, this parameter qualified five patients (three from the wide pQRS and two from the narrow pQRS groups) as suboptimal LBBAP (IVDS was also negative in all five patients).

Identifying a classic ECG parameter capable of predicting TVACT was less clear as, due to variations in anatomy and patch placement, a narrow pQRS cannot be the only predictor of good TVACT. As demonstrated by our data (Figure 5 / Table 5), while there was a 100% correlation between pQRS and TVACT in the narrow pQRS group, the wide pQRS group exhibited some discrepancies. After analysis, another 12-lead ECG parameter – dRVAT, could be used in addition to pQRS and V1AD to predict a good TVACT. Our study suggests that when dRVAT > 120 ms it corresponded to a very long TVACT and suboptimal ventricle activation; however, if this parameter was < 120 ms, the activation was close to physiological and might be accepted.

### Intrinsic vs. paced

Comparison with the intrinsic rhythm was also an important part of our research as we could assess the physiological nature of the pacing in a particular patient in terms of intraventricular synchrony and bundle branch block correction (if present).

In our study, we categorized our patients into two groups—narrow and wide intrinsic QRS—and analyzed both intrinsic and paced rhythms using ECGI. The main criterion was intraventricular dyssynchrony represented by the IVDS value (Table 5). Our study confirms that LBBAP can preserve ventricle activation times in patients with initially narrow QRS while significantly improving it in a native wide-QRS cohort, in agreement with other investigators. (14, 15)

### Suggested implantation workflow

Based on our study, we recommend adding extra steps to intra- and postprocedural LBBAP confirmation by analyzing two extra parameters – V1AD and dRVAT, which can potentially help to identify physiological pacing without using ECGI. The suggested implantation workflow is summarized in Figure 5.

The long-term effect of intraventricular synchrony/dyssynchrony after LBBAP device implantation needs to be assessed in future studies.

### Limitations

This study reports a single-center experience. Furthermore, it is a nonrandomized trial conducted in a relatively limited number of patients. Heart failure patients were excluded from the study. There were no special échographic protocol used to asses intraventricular synchrony.

Two patients from the narrow pQRS group were found to present significant ventricular dyssynchrony despite correct ECG parameters; they have been programmed for follow-up to confirm our findings as this could potentially cause hemodynamic issues.

## 5. Conclusions

ECGI can bring significant value to assessing the efficacy of new pacing modalities and provide a greater amount of data for the precise determination of implantation outcomes, including detailed activation assessment and comparison with intrinsic conduction. Key ECGI values confirming proper ventricular activation were defined, and the corresponding 12-lead parameters were identified which may help to predict ventricular synchrony using 12 lead ECG only during implantation. Further study is required to determine whether classical LBBAP implantation criteria and modality classification need to be updated based on this new information.

## Funding

This research received no external funding.

## Institutional Review Board Statement

The study was conducted in accordance with the Declaration of Helsinki and approved by the Ethics Committee of Universitair Ziekenhuis Brussel.

## Informed Consent Statement

Informed consent was obtained from all subjects involved in the study.

## Data Availability Statement

The data that support the findings of this study are available from the corresponding author upon reasonable request.

## Conflicts of Interest

Dr Almorad has received instituitionnal research grants and compensation for teaching from Biosense Webster, Medtronic, Boston Scientific. Dr. de Asmundis has received research grants on behalf of the center from Biotronik, Medtronic, Abbott, LivaNova, Boston Scientific, AtriCure, Philips, and Acutus; and has received compensation for teaching purposes and proctoring from Medtronic, Abbott, Biotronik, Livanova, Boston Scientific, Atricure, and Acutus Medical Daiichi Sankyo. Dr. Chierchia has received compensation for teaching purposes and proctoring from Medtronic, Abbott, Biotronik, Boston Scientific, and Acutus Medical. Dr. La Meir has been a consultant for Atricure. Mr Zeng and Dr. Eltsov are receiving compensations from Medtronic.

## List of Abbreviations

LBB: Left Bundle Branch
LBBB: Left Bundle Branch Block
LBBP: Left Bundle Branch Pacing
LBFP: Left Bundle Fascicular Pacing
LBBAP: Left Bundle Branch Area Pacing
LVSP: Left Ventricular Septal Pacing
RVSP: Right Ventricular Septal Pacing
HBP: His bundle pacing
CSP: Conduction System Pacing
RV: Right Ventricle
LV: Left Ventricle
LVAT: Left Ventricle Activation Time (ECG)
RVAT: Right Ventricle Activation Time (ECG)
V1AD: V1-V6 Activation Delay
PM: Pacemaker
EAM: Electroanatomical Mapping System
ECGI: Electrocardiographic Imaging
GA: General Anesthesia
EGM: Electrogram
TVACT: Total ventricular activation time (ECGI)
LVACT: Left Ventricle Activation Time (ECGI)
LVACTi: Intrinsic LVACT (ECGI)
RVACT: Right Ventricle Activation Time (ECGI)
RVACTi: Intrinsic RVACT (ECGI)
IVDS: Intraventricular Deisynchrony

## Notes

### Competing Interest Statement

Dr Almorad receives compensation for teaching from Biosense Webster, Dr. de Asmundis has received research grants on behalf of the center from Biotronik, Medtronic, Abbott, LivaNova, Boston Scientific, AtriCure, Philips, and Acutus; and has received compensation for teaching purposes and proctoring from Medtronic, Abbott, Biotronik, Livanova, Boston Scientific, Atricure, and Acutus Medical Daiichi Sankyo. Dr. Chierchia has received compensation for teaching purposes and proctoring from Medtronic, Abbott, Biotronik, Boston Scientific, and Acutus Medical. Dr. La Meir has been a consultant for Atricure. Mr Zeng and Dr. Eltsov are receiving compensations from Medtronic.

### Clinical Trial

NCT05401851

### Author Declarations

The study had been approved by ethics committee of University Hospital of Brussels (UZ Brussel)

